# SeroTracker-RoB: a decision rule-based algorithm for reproducible risk of bias assessment of seroprevalence studies

**DOI:** 10.1101/2021.11.17.21266471

**Authors:** Niklas Bobrovitz, Kim Noël, Zihan Li, Christian Cao, Gabriel Deveaux, Anabel Selemon, David A. Clifton, Mercedes Yanes-Lane, Tingting Yan, Rahul K. Arora

## Abstract

**Background:** Risk of bias (RoB) assessments are a core element of evidence synthesis but can be time consuming and subjective. We aimed to develop a decision rule-based algorithm for RoB assessment of seroprevalence studies.

**Methods:** We developed the SeroTracker-RoB algorithm. The algorithm derives seven objective and two subjective critical appraisal items from the Joanna Briggs Institute Critical Appraisal Checklist for Prevalence studies and implements decision rules that determine study risk of bias based on the items. Decision rules were validated using the SeroTracker seroprevalence study database, which included non-algorithmic RoB judgements from two reviewers. We quantified efficiency as the mean difference in time for the algorithmic and non-algorithmic assessments of 80 randomly selected articles, coverage as the proportion of studies where the decision rules yielded an assessment, and reliability using intraclass correlations comparing algorithmic and non-algorithmic assessments for 2,070 articles.

**Results:** A set of decision rules with 61 branches was developed using responses to the nine critical appraisal items. The algorithmic approach was faster than non-algorithmic assessment (mean reduction 2.32 minutes [SD 1.09] per article), classified 100% (n=2,070) of studies, and had good reliability compared to non-algorithmic assessment (ICC 0.77, 95% CI 0.74-0.80). We built the SeroTracker-RoB Excel Tool which embeds this algorithm for use by other researchers.

**Conclusions:** The SeroTracker-RoB decision-rule based algorithm was faster than non-algorithmic assessment with complete coverage and good reliability. This algorithm enabled rapid, transparent, and reproducible RoB evaluations of seroprevalence studies and may support evidence synthesis efforts during future disease outbreaks. This decision rule-based approach could be applied to other types of prevalence studies.

## INTRODUCTION

Seroprevalence studies use antibody tests to estimate the prevalence of infection or vaccination.^1^ These studies have been used for decades to measure the true extent of infection,^2^ quantify protection resulting from previous infection or vaccination, and inform public health measures.

During the COVID-19 pandemic, there has been an unprecedented increase in the utilization of seroprevalence studies, with results reported from over 3,500 such studies as of June 1, 2022.^3^ These studies have made important contributions to the pandemic response, but their methods have varied widely.^2,4^ Accordingly, robust risk of bias (RoB) assessments have been crucial for synthesizing and utilizing trustworthy seroprevalence data for public health decision-making.

Risk of bias refers to systematic error in study results that can arise because of flaws in study design, conduct, analysis, interpretation, or reporting. As such, risk of bias assessment includes identification of study factors and safeguards that protect against systematic error and an empirical construct for making a judgement about the potential bias resulting from the absence of these factors and safeguards (e.g., low, moderate, high risk of bias).^5^ These constructs vary but can include counting the number of missing study safeguards or identifying important patterns of missing safeguards, depending on the assessors’ judgements regarding the possible influence on study estimates.

There are several validated tools for RoB assessment of prevalence studies but little consistency in the use of these tools across evidence synthesis efforts.^6,7^ This demonstrates uncertainty about the most appropriate tool for use in evidence synthesis. Furthermore, the validated tools available include heterogeneous evaluation criteria, are time-consuming to use, and yield RoB assessments with inherent variation given a reliance on evaluators’ subjective judgements.^8-11^ These limitations present a major challenge for rapid and reproducible synthesis of seroprevalence studies. Indeed, meta-epidemiological reviews of prevalence studies suggest that a unified approach to RoB assessment using a validated tool is needed.^6^

The Joanna Briggs Institute (JBI) Critical Appraisal Checklist for Prevalence Studies is an appropriate candidate to form the basis for such a unified approach.^6,7^ The tool was developed for the explicit purpose of appraising disease prevalence studies by the Joanna Briggs Institute, an organization that develops methodologies and guidance on the process of conducting systematic reviews.^12^ Items were developed based on systematic reviews of prevalence assessment and expert opinion to address key issues of internal and external validity for prevalence studies (i.e., presence or absence of key study safeguards against systematic error).^13^ The tool has been validated by multiple investigator groups and compares favorably to other tools in terms of content validity, construct validity, and usability.^6,14,15^ Furthermore, the JBI tool is the most commonly utilized tool for appraisal of prevalence studies in evidence synthesis projects, indicating a greater level of support for this tool relative to the many others available. However, the nine-item JBI checklist requires subjective judgement for completion. This tool also has no provision for overall RoB assessment, which is often sought by investigators as it can serve as a valuable summary metric for analysis, reporting, and interpretation of a collection of evidence (e.g., sub-group analysis that exclude studies at higher risk of bias). As a result, this checklist has been operationalized differently by groups conducting evidence synthesis, with each effort defining their own approach to derive overall RoB assessments.^16-19^ In some instances, this has resulted in markedly different item ratings and RoB assessments for the same body of underlying literature.^16,19^

We aimed to develop an objective, rapid, and reproducible approach to RoB assessment for seroprevalence studies. In this manuscript, we describe the SeroTracker-RoB decision rule-based algorithmic, which involves use of a critical appraisal tool derived from the JBI checklist and application of decision rules to the critical appraisal results to yield an overall RoB assessment. We evaluated the efficiency, coverage, and reliability of this algorithmic approach, and developed an Excel tool that implements this algorithm for use by other researchers.

## METHODS

### The SeroTracker-RoB decision rule-based algorithm

We developed an algorithm for RoB assessments of seroprevalence that involves two components: (1) a critical appraisal checklist that was derived from the Joanna Briggs Institute Critical Appraisal Checklist for Prevalence Studies^12^; (2) decision rules that are applied to the responses of the critical appraisal checklist.

#### Component 1: Critical appraisal checklist derived from the JBI checklist

The critical appraisal checklist we use is an operationalized version of the JBI Critical Appraisal Checklist for Prevalence Studies. Our operationalization of the items aimed to reduce the number of subjective judgements required and instead allow objective completion based on routinely extracted data from a given seroprevalence study.^20^

The original JBI checklist requires that reviewers subjectively judging nine categorical items based on data reported in a prevalence study: (1) sample frame appropriateness; (2) sampling method; (3) sample size/calculation; (4) subject and setting described in detail; (5) representativeness of sample within analysis; (6) test sensitivity and specificity; (7) consistent test use; (8) appropriate statistical adjustment; and (9) response rate.^12^

In our operationalization, six of the nine JBI checklist items (item 2, 3, 5, 6, 7, 8) were modified to add conditions that enabled them to be judged in a binary fashion based on objective data reported in a prevalence study (Table 1, Supplementary File 1). For example, Item 2 (were participants recruited in an appropriate way?) was marked as “Yes” if probability sampling methods or entire population sampling was used and “No” if any other method was used.

**Table 1.**
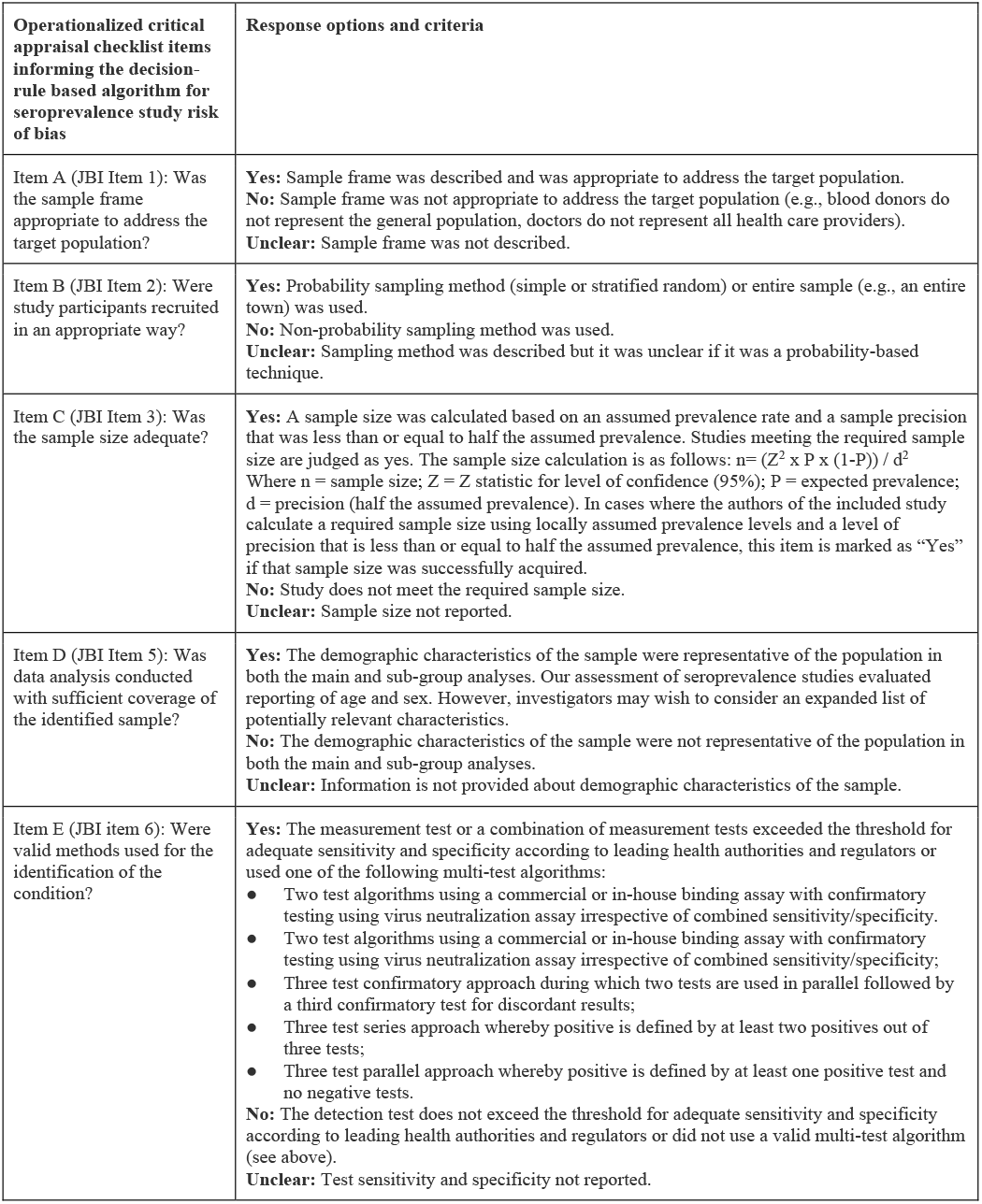
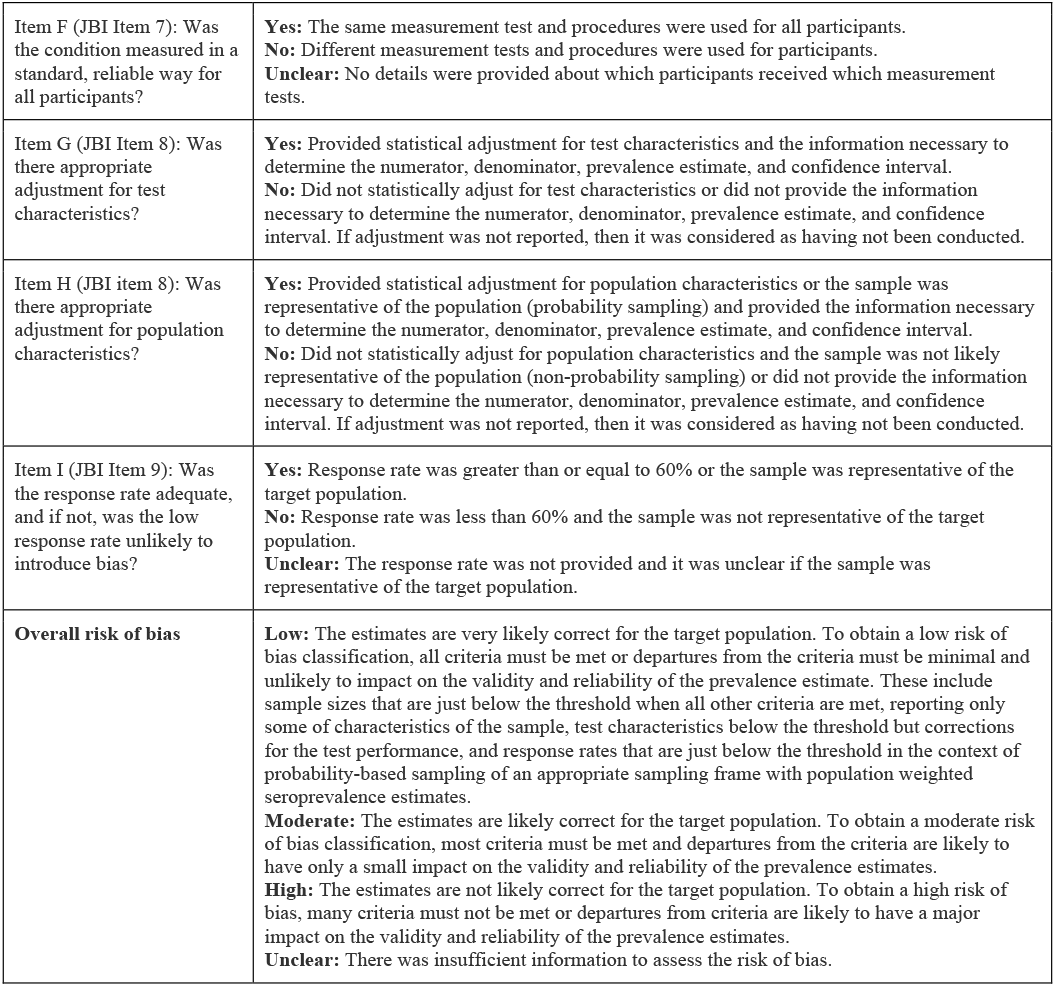
Summary of operationalized critical appraisal checklist items derived from the Joanna Briggs Institute Checklist for Prevalence Studies.

Item 8 (appropriate statistical analysis) was split in two parts to differentiate studies that adjusted prevalence estimates for test sensitivity and specificity but not population demographics and vice versa. We separated these items as failing to adjust for test characteristics and failing to adjust for population characteristics have different risk of bias implications from one another. Item 4 was eliminated as it is a reporting item that would not impact on risk of bias. Full details of the operationalization for each item are reported in Supplementary File 1.

To clarify when we are referring to the operationalized version and original version, we relabelled the operationalized items using alphabetic characters (Item A-J) and will hereafter refer to the operationalized checklist as the critical appraisal checklist (Supplementary File 1).

The critical appraisal checklist items were then categorized as relating to selection biases or measurement biases. Selection biases related to the extent to which the seroprevalence estimated in the study sample was representative of the seroprevalence in the target population (items A, B, C, D, H, I). Measurement biases were those related to measurement error (items E, F, G).

As a first step towards a tool to implement the algorithmic RoB evaluations, we created coding logic for each critical appraisal item based on key information that would be extracted as part of a critical appraisal or systematic review of prevalence studies (Supplementary File 1).^21^ Seven of the nine items can be instantaneously completed using this coding logic applied to data extracted from studies. Coding logic could not be created for two items, as they require study-, sample-, and target population-specific contextual judgements: item A (whether the sample frame was representative of the target population) and item D (whether the characteristics of the sample were representative of the target population, in both the main and sub-group analyses). We added to pre-existing JBI guidance to offer detailed instructions and illustrative examples on how to complete these two subjective items.

#### Component 2: Developing decision rules for risk of bias assessment

We developed decision rules that could be applied to the critical appraisal items responses to generate an overall RoB assessment (low, moderate, or high) for each study (Figure 1). The use of decision rules was selected as it allows for all checklist items to be considered together in evaluating overall study RoB. The decision rules were developed based on published guidance on estimating disease prevalence^21^, reports on the evaluation of prevalence studies^6,7,22^, evidence of the impact of bias on research results^23-28^, opinions of experts in evidence synthesis and infectious disease epidemiology, and guidance created by SeroTracker researchers after evaluation of thousands of seroprevalence studies (Supplementary File 1).^2-4^

**Figure 1.**
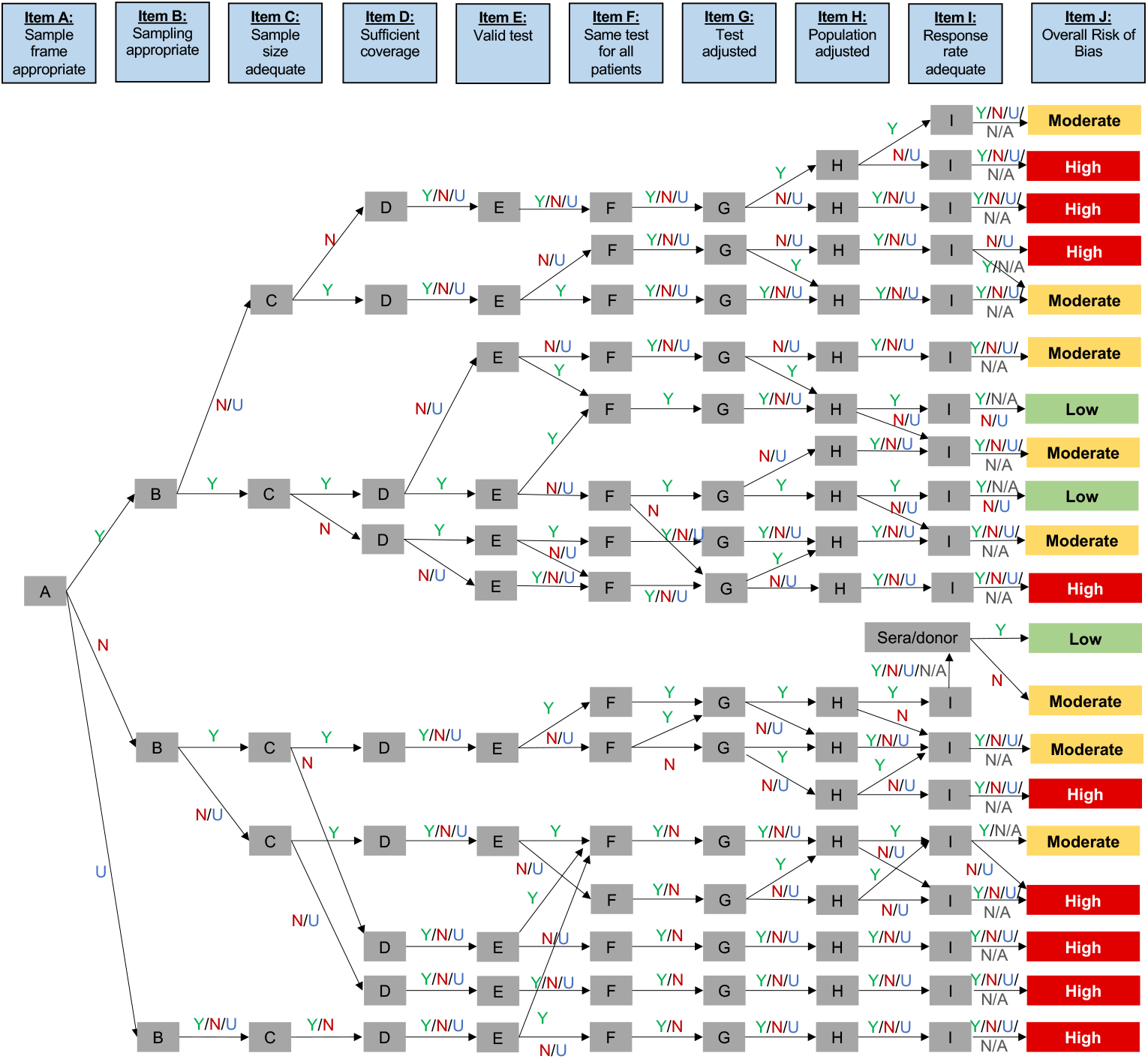
SeroTracker Decision Rules for Risk of Bias Assessment

The decision rules were based on the extent of selection and measurement bias present in a study. Meta-epidemiological studies have shown that selection and measurement biases have been associated with overestimates and underestimates.^23-28^ As such, the extent of these biases was mapped to the different levels of risk of bias.

We defined selection bias to occur when the sample from which estimates are derived differs systematically from the target population. Selection bias may arise from factors affecting sample recruitment, size, and retention including using a sample frame that is not representative of the target population, a low response rate which may increase the chance that participants differ from non-participants and drop-outs, and a low sample size which may increase the chance of known and unknown imbalances in participant characteristics compared to the target population.^22,25,26^ We defined measurement bias to occur when there is systematic error in the measurement of the outcome. Measurement bias may arise from low detection test sensitivity and specificity or use of different measurement methods for different participants.^22,27,28^

The decision rules considered studies to be low RoB if there was no selection bias (i.e., appropriate sample frame, probability sampling, adequate sample size, and statistical adjustment for population characteristics) and no measurement bias (i.e., adequate sensitivity/specificity and/or adjustment for test performance). These rules generally considered studies at moderate RoB if they had some selection bias (e.g., inappropriate sample frame but probability sampling, adequate sample size, and statistical adjustment for population characteristics) or no measurement biases. In contrast, studies were considered high RoB if the study had considerable selection bias (e.g., non-probability sampling and inadequate sample size) and considerable measurement bias (i.e., poor sensitivity/specificity and lack of adjustment for test performance).

### Validation dataset

The SeroTracker-RoB decision-rule based algorithm was evaluated using data from SeroTracker’s living systematic review database of SARS-CoV-2 seroprevalence studies.^3^ The protocol for the review was registered and published (PROSPERO: CRD42020183634, version July 21, 2021).

The dataset used for validation included all seroprevalence studies included in the SeroTracker database with publication dates between January 1, 2020 to November 17, 2021. The 2,070 studies in this evaluation dataset included peer-reviewed literature, preprints, government and non-governmental organization reports, and media articles.

Non-algorithmic RoB and algorithmic RoB assessments were completed for each study. Non-algorithmic RoB assessments were completed by two independent reviewers, each of whom examined data routinely extracted from each article to complete the critical appraisal checklist (Supplementary File 1) and performed a RoB assessment using general guidance that we developed and used as a team prior to the development of the algorithm (Supplementary File 1). This guidance described key considerations for assessing risk of bias but did not function as a formalized algorithm. There were five directives regarding conditions wherein studies should not be rated as low risk of bias; however, we did not measure adherence to these directives and reviewers could override the guidance if they provided reasonable justification and a second reviewer agreed with the justification.

Disagreements were resolved by consensus. Algorithmic RoB assessments were conducted using the SeroTracker-RoB decision rule-based algorithm as outlined above, involving application of the coding logic for the critical appraisal checklist items and decision rules to the same data that was used to generate the non-algorithmic assessments. The coding logic and decision rules were implemented in an AirTable programmable spreadsheet.

### Evaluating efficiency, coverage, and reliability

#### Sample size

We calculated the required sample size for analysis to precisely estimate inter-rater reliability between the algorithmic and non-algorithmic approaches.^29-32^ To inform the sample size calculation an estimate of the expected reliability was required. We used estimates of reliability for the non-algorithmic approach, the baseline approach to RoB assessment for articles included in the SeroTracker database, to serve as the expected reliability in the calculation. The database included ratings for 2,070 articles conducted by 12 reviewers in pairs. The reliability for the non-algorithmic approach was determined by calculating absolute agreement using a two-way random-effects single-measures ICC and was found to be 0.74 (95% CI 0.71-0.76). With k=2 raters (non-algorithmic vs. algorithm), alpha=0.05, and an anticipated ICC of 0.74 the required sample size to estimate ICC within a precision of 0.2 was 80.

#### Efficiency

We assessed efficiency by comparing time to complete non-algorithmic and algorithmic RoB assessments for 80 seroprevalence articles that were randomly selected from the SeroTracker dataset. Five SeroTracker systematic reviewers were timed as they each completed between five and fifty non-algorithmic assessments. The time required for the non-algorithmic approach included assessment of each checklist item and evaluation of overall RoB. The time required for the algorithmic approach included only the time taken to evaluate two critical appraisal checklist items requiring subjective judgement (Item A, Item D). There was instantaneous application of the coding logic for the other seven objectively determined critical appraisal items and the decision rules for RoB. We quantify the mean difference in time between the non-algorithmic and algorithmic approaches. Formal statistical testing was not conducted as all algorithmic assessments were, by definition, shorter in duration given their instantaneous completion based on coding logic.

#### Coverage

To determine coverage, we calculated the proportion of studies in the dataset for which the decision rule-based algorithm yielded a RoB assessment.

#### Reliability

To evaluate reliability of the RoB assessments across consensus non-algorithmic and algorithmic approaches, we calculated the absolute agreement between ordinal overall RoB assessments using a two-way random-effects average-measures intraclass correlation (ICC).^29-31^ We used the entire SeroTracker database of 2,070 articles to assess the reliability of the algorithmic compared to non-algorithmic approach. All data are available at SeroTracker.com. Analyses were conducted using STATA 14 (College Station, TX: StataCorp LP).

## RESULTS

### Decision rules for RoB

The decision tree to classify overall study RoB is shown in Figure 1. The decision tree classified overall RoB as low, moderate, or high based on the categorical ratings for each critical appraisal checklist item, considering each item in turn to arrive at an overall judgement (i.e., item A, item B, etc.). The tree included 61 decision rules, which could lead to low (n=6), moderate (n=29), or high (n=26) RoB.

### Efficiency

Use of the SeroTracker-RoB algorithmic approach resulted in faster RoB assessments compared to the non-algorithmic approach with a mean reduction of 2.32 minutes (SD 1.09) per article (algorithm mean 0.64 [SD 0.24] vs. non-algorithm mean 2.97 [SD 1.16]). The algorithmic approach took less time than the non-algorithmic approach for all 80 articles evaluated.

### Coverage

The algorithmic approach yielded a RoB assessment for 100% (n = 2,070) of the SARS-CoV-2 seroprevalence studies in the SeroTracker database.

### Reliability

ICC for the reliability of the RoB assessment was 0.77 (95% confidence interval [CI] 0.74-0.80) between the SeroTracker-RoB algorithmic approach and consensus non-algorithmic review. A summary of the risk of bias assessments for the algorithmic and non-algorithmic approaches are shown in Figure 2.

**Figure 2.**
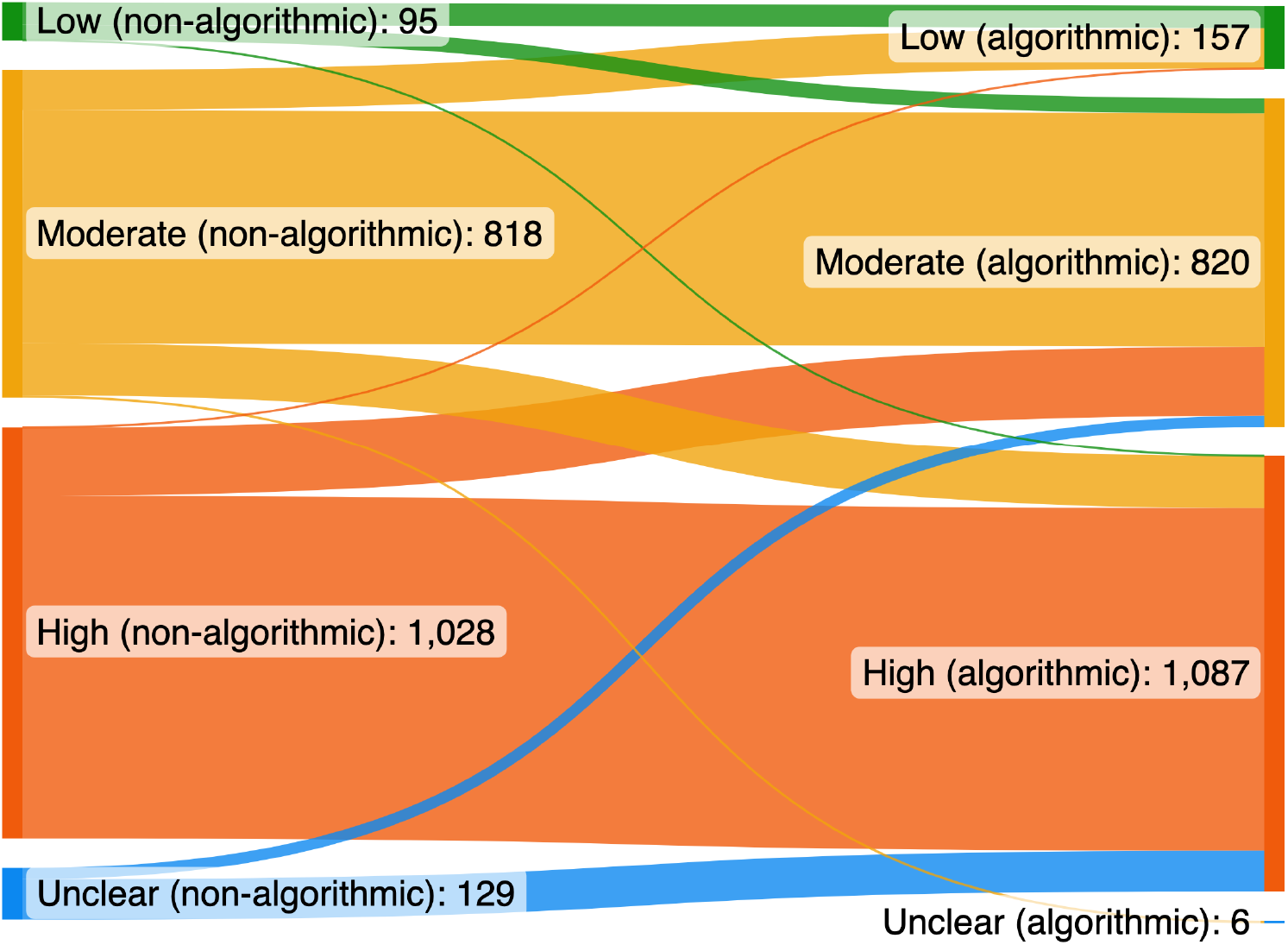
Comparison of non-algorithmic and algorithmic risk of bias assessments for 2,070 seroprevalence studies

### The SeroTracker-RoB Excel Tool

A Microsoft Excel tool (the “SeroTracker-RoB Excel Tool”) that implements the algorithmic approach to prevalence study RoB assessment can be found in Supplementary File 2. This tool includes the operationalized critical appraisal checklist items with coding logic and decision rules. The decision tree logic was implemented in Excel using Visual Basic for Applications.

The SeroTracker-RoB Excel Tool includes four sheets: (1) a legend sheet to describe the tool and orient the user; (2) a data extraction sheet with 25 core data fields for reviews of prevalence studies, including the two critical appraisal checklist items requiring subjective assessment; seven user-defined thresholds for objectively determined critical appraisal items (i.e., minimum sample size, test validity, response rate); seven objective critical appraisal item variables; and a variable with the algorithmically determined RoB rating; (3) a data dictionary sheet with descriptions of the data extraction fields, instructions for extraction, and the critical appraisal item criteria and coding logic for each item; and (4) a data validation sheet where options for dropdown menus can be edited by the user. Using built in formulas and these extracted fields, the tool completes the checklist and produces an overall RoB rating. Users who wish to manually enter responses to the critical appraisal items, as opposed to rely on coding logic applied to extracted study data, can enter their ratings into the nine critical appraisal item variables, and still take advantage of the SeroTracker-RoB decision rules for an algorithmically determined overall RoB assessment.

## DISCUSSION

We developed the SeroTracker-RoB decision rule-based algorithm for RoB assessments of seroprevalence studies. This tool has the potential to lay the foundation for rapid, robust, and reproducible RoB assessment of prevalence studies.

There is no consensus on the most valid approach to expediting RoB assessments.^6,7^ Some studies have used summary scores, adding the number of criteria met for a given critical appraisal checklist and creating a threshold on that score to assess RoB.^16-18^ However, this approach weights each checklist item equally, which does not reflect the different implications of critical appraisal concepts for RoB. For example, when considering different item ratings for the critical appraisal checklist, marking six out of nine items “Yes” could be achieved by 84 different combinations of item responses, which have different implications for RoB.^22,25-28,33-35^

A comparison of two reviews of seroprevalence studies in Africa highlights how different approaches to risk of bias based on the JBI checklist can result in different assessments for the same underlying body of literature.^16,19^ The reviews used similar search strategies and study inclusion criteria. One review used a summary scoring method whereby more JBI items scored as “Yes” indicated lower risk of bias.^16^ The items rated as “Yes” were summated and 9, 5–7 and ≤4 indicated high quality, moderate quality and poor quality studies. They found that the majority were generally at lower risk of bias (74% high quality, 22% moderate quality, 4% low quality). In contrast a similar review using the SeroTracker algorithmic approach found that most studies were at moderate or high risk of bias (24% low risk of bias, 40% moderate risk of bias, 37% high risk of bias).^19^

Weighted averages better reflect the relevance of each item, overcoming some limitations of simple summary scores. However, checklist items cannot always be considered independently in assessing RoB, thereby introducing complexity in the derivation of a weighted score. For example, it is more important to correct for antibody test performance (item I = yes) when assay sensitivity and specificity are low (item E = no).

Considering combinations of items together may enable better expedited RoB assessment. Some studies have trained deep learning algorithms to identify relevant text in publications of randomized controlled trials and predict RoB assessments, with reasonable accuracy compared to human reviewers.^33,34^ However, these algorithms have largely been trained on small datasets and have limited interpretability and transparency due to the “black-box” decision making of the models.

A decision rule-based approach, on the other hand, provides a transparent and interpretable model to expedite RoB assessment.^35^ The tree structure captures interactions between critical appraisal checklist features, is easy to implement, and enables clear visualization of which combinations of features are most important for RoB. The decision tree in the SeroTracker-RoB algorithmic approach reveals two key axes of bias in seroprevalence studies: selection biases, related to the correspondence between the sample frame and target population, sampling representativeness, response rate, and population weighting; and measurement biases, related to detection test performance, correction for that performance, and consistency of test use across participants. Clusters of rules in this decision tree also make clear how features in each axis interact to determine study RoB.

RoB assessment is a time-consuming component of conducting systematic reviews.^5,8^ As such, improving efficiency of the assessment process may help to reduce the burden of evidence synthesis. This is particularly important for living reviews and reviews during health emergencies, which may need to process a high volume of information quickly to inform public health decision making.

The SeroTracker project exemplifies how time-savings can be of value. There is an average of 35 new studies added each week to the SeroTracker living systematic review project.^3^ The SeroTracker-RoB algorithmic approach described in this manuscript is 4.6 times faster than the non-algorithmic approach for assessing study RoB, which took approximately three minutes. Thus, using the algorithmic approach would save nearly three hours of reviewer time each week.

In traditional non-algorithmic RoB assessment, there is imperfect agreement between even trained human reviewers.^5,8-10^ Inter-rater reliability for RoB between two independent reviewers using the non-algorithmic approach was moderate (ICC 0.74). As such, non-algorithmic review cannot be considered a perfect standard, and the 0.77 ICC between the SeroTracker-RoB algorithmic and non-algorithmic approach in part reflects the heterogeneity and inconsistency of non-algorithmic assessment. This highlights the benefit of an algorithmic approach that yields reproducible assessments.

There have been many systematic reviews of severe acute respiratory syndrome coronavirus 2 (SARS-CoV-2) seroprevalence studies to date.^2,4,16-18,36^ Several of these reviews use the JBI checklist as a foundation for critical appraisal, but they implement the checklist and judge overall RoB in different ways.^2,4,16-18^ For this reason, we developed the SeroTracker-RoB Excel Tool (Supplementary File 2), which embeds the coding logic for instantaneous completion of seven of the nine critical appraisal items and application of the decision rules in a user-friendly data extraction sheet. This tool may be valuable to other investigators seeking to conduct rapid and reliable RoB assessments for prevalence studies — an important endeavor, given the ongoing value of SARS-CoV-2 seroprevalence studies for surveillance as the virus becomes endemic^37^, and given the many prevalence studies conducted for other diseases and conditions.

The decision rule-based algorithm may be suitable for use in evaluating a broad array of prevalence studies. Users of the algorithm tool can adapt the thresholds for the objectively determined critical appraisal items to meet their unique needs. For example, we judged valid methods of detecting SARS-CoV-2 antibodies using 90% sensitivity and 97% specificity thresholds established by the World Health Organization^38^, but this criterion will vary for other conditions. To accommodate these needs, the SeroTracker-RoB Excel Tool includes embedded user-set thresholds that can easily be altered as required. Our description of each item in Supplementary File 1 also includes clear instructions on which criteria users may wish to alter for their own purposes.

This study had several strengths. Firstly, to our knowledge, this is the first validation of decision rules for RoB in seroprevalence studies, with the validation dataset including thousands of heterogeneous SARS-CoV-2 seroprevalence studies across study designs, regions, and target populations. Second, the use of a decision rule approach enables transparent RoB assessment, while also ensuring reproducibility and speed. Finally, the decision rules have a robust foundation in the validated and widely used JBI critical appraisal checklist, published guidance on estimating disease prevalence, evidence on the impact of bias on empirical estimates, opinions of methodological experts, and the experiences of researchers that have conducted thousands of RoB assessments for seroprevalence studies.

This study had several limitations. First, the SeroTracker-RoB algorithmic approach requires reviewers to make subjective judgements on two critical appraisal items (Item A, Item D), which may be associated with response variation and reviewer burden. However, we provide clear guidance for making those assessments. It is theoretically possible to develop objective standards for the completion of Item A and Item D however, there are practical challenges to this. For example, Item A (Was the sample frame appropriate to address the target population?) could be operationalized by determining whether evidence had been cited to show that the proposed sample frame had previously been shown to be representative of a target population. However, this would require substantial pre-existing literature which may not exist for every topic. Item D (whether the characteristics of the sample were representative of the target population, in both the main and sub-group analyses) could be operationalized by assessing whether statistical testing demonstrated that the sample demographics were not different from the population demographics for every analysis. However, this would still require judgement about which demographic characteristics should be matching. The relevant set of characteristics may be specific to the diseases and populations being studied. Thus, subjectivity and content expertise may always be required for these items. Second, the decision rule was derived in part using expert judgement. However, the transparency of this algorithm allows for scientific debate and further refinement of the decision rules, if needed. Thirdly, the generalizability of the SeroTracker-RoB decision rules is unclear. Although the validation database used was large and robust, the data came exclusively fromSARS-CoV-2 seroprevalence studies. Applying this approach to prevalence studies for other pathogens or conditions may require additional validation or adaptation.

## Conclusions

We developed and validated the SeroTracker-RoB decision rule-based algorithm, which enables rapid, transparent, and reproducible risk of bias assessment for seroprevalence studies. This algorithm includes seven objectively determined critical appraisal items and two subjectively determined critical appraisal items derived from the Joanna Briggs Institute Critical Appraisal Checklist for Prevalence studies, and interpretable decision rules to assess overall risk of bias.

The SeroTracker-RoB Excel Tool embeds the algorithm in a data extraction sheet enabling its use by other researchers conducting evidence synthesis of seroprevalence studies and adaptation by researchers conducting evidence synthesis of other types of prevalence studies.

## Data Availability

All data produced are available online at SeroTracker.com

https://serotracker.com/

## What is already known

Risk of bias assessments are a core element of evidence synthesis but can be time consuming and subjective. As such, there is a need to expedite objective and reproducible assessments using validated tools. Rapid evaluation of infection prevalence studies is of particular importance given that these studies are conducted during disease outbreaks and pandemics to inform public health decision making. However, there are currently no tools for expedited risk of bias assessment of prevalence studies.

## What is new

We developed a reproducible algorithmic approach to risk of bias assessment for SARS-CoV-2 seroprevalence studies. This algorithm includes seven objectively determined critical appraisal items and two subjectively determined critical appraisal items derived from the Joanna Briggs Institute Critical Appraisal Checklist for Prevalence studies, and a decision tree which determines risk of bias based on checklist items. The algorithmic approach was 4.6 times faster than traditional non-algorithmic assessment, successfully categorized all 2,070 studies that it was tested on, and had good agreement with non-algorithmic assessments. We built a simple Excel tool so that other researchers can use this algorithmic approach.

## Potential impact for Research Synthesis Methods readers outside the authors’ field

The SeroTracker-RoB decision rule-based algorithm and Excel Tool enable rapid, transparent, and reproducible risk of bias assessments for SARS-CoV-2 seroprevalence studies and could be readily adapted for other types of prevalence studies. Moreover, we show how to operationalize a critical appraisal checklist and develop a decision tree to algorithmically generate risk of bias assessments. These processes may be applicable to risk of bias assessment for other types of studies and in other scientific disciplines.

