## Supplementary File 1 for "SeroTracker-RoB: a decision rule-based algorithm for reproducible risk of bias assessment of seroprevalence studies"

**Supplementary file 1. Full details of the critical appraisal checklist items derived from the Joanna Briggs Institute Checklist for Prevalence Studies and principles guiding the non-algorithmic and algorithmic risk of bias assessment**

| **Item A (JBI Item 1): Was the sample frame appropriate to address the target population?** | |
|---|---|
| Response: Yes | Sample frame was described and was appropriate to address the target population. |
| Response: No | Sample frame was not appropriate to address the target population (e.g., blood donors do not represent the general population, doctors do not represent all health care providers). |
| Response: Unclear | Sample frame was not described. |
| Notes | |
| Rationale for operationalization | This item requires subjective evaluation as to whether the sample frame appropriately matches the target population. JBI guidance reports the following instructions for subjective evaluation of this item: "the term "target population" should not be taken to infer every individual from everywhere or with similar disease or exposure characteristics. Instead, give consideration to specific population characteristics in the study, including age range, gender, morbidities, medications, and other potentially influential factors. For example, a sample frame may not be appropriate to address the target population if a certain group has been used (such as those working for one organization, or one profession) and the results then inferred to the target population (i.e., working adults). A sample frame may be appropriate when it includes almost all the members of the target population (i.e., a census, or a complete list of participants or complete registry data)."[1] |
| Coding logic: *variables* (outcome) | Not applicable. |
| **Risk of bias domain** | Selection bias. |
| Principles guiding non-algorithmic risk of bias assessment and used to inform development of the algorithm | If this item is "No" then the study will not be marked at low risk of bias. There is only one borderline case: blood donor or residual sera studies with all other items met, including very large sample sizes, and very robust population and test adjustment. |

<table>
<tr><td colspan="2">Item B (JBI Item 2): Were study participants recruited in an appropriate way?</td></tr>
<tr><td>Response: Yes</td><td>Probability sampling method (simple or stratified random) or entire sample (e.g., an entire town) was used.</td></tr>
<tr><td>Response: No</td><td>Non-probability sampling method was used.</td></tr>
<tr><td>Response: Unclear</td><td>Sampling method was described but it was unclear if it was a probability-based technique.</td></tr>
<tr><td colspan="2">Notes</td></tr>
<tr><td>Rationale for operationalization</td><td>The rigor of sampling methods is indicative of the risk of selection bias. The two sampling methods at lowest risk of selection bias are probability-based sampling and use of the entire sample frame. As such, these two methods were selected to represent adequate sampling. A binary judgement was created using these sampling methods to reduce decision burden.<br><br>If the sampling method is not described at all then investigators should consider excluding the study as sampling method is a major consideration for selection bias.</td></tr>
<tr><td>Coding logic: variables (outcome)</td><td>This item may be completed using a single variable:<br>● Sample method - a categorical variable describing the type of sampling method<br><br>This one variable (outcome) determines Item B (outcome) as follows:<br>● Sample method (Blank) = Item B (Missing)<br>● Sample method (Unclear) = Item B (Unclear)<br>● Sample method (Simple random) = Item B (Yes)<br>● Sample method (Stratified random) = Item B (Yes)<br>● Sample method (Entire sample) = Item B (Yes)<br>● Sample method (All other methods*) = Item B (No)<br><br>*Users of this tool may wish to specifically label other methods (e.g., convenience sample).</td></tr>
<tr><td>Risk of bias domain</td><td>Selection bias.</td></tr>
<tr><td>Principles guiding non-algorithmic risk of bias assessment and used to inform development of the algorithm</td><td>If this is “No” then the study will not be marked at low risk of bias.</td></tr>
</table>

| **Item C (JBI Item 3): Was the sample size adequate?** | |
|---|---|
| Response: Yes | A sample size was calculated based on an assumed prevalence rate and a sample precision that was less than or equal to half the assumed prevalence. Studies meeting the required sample size are judged as yes. The sample size calculation is as follows:<br><br>$n = (Z^2 \times P \times (1-P)) / d^2$<br><br>Where n = sample size<br>Z = Z statistic for level of confidence (95%)<br>P = expected prevalence<br>d = precision (half the assumed prevalence)<br><br>In cases where the authors of the included study calculate a required sample size using locally assumed prevalence levels and a level of precision that is less than or equal to half the assumed prevalence, this item is marked as "Yes" if that sample size was successfully acquired.<br><br>To calculate the required sample size for evaluation of our sample of seroprevalence studies we used an assumed prevalence of 2.5%, which was the global average estimated by the WHO in April, 2020.[2] Based on guidance by the Joanna Briggs Institute and published medical statistical recommendations we selected a precision value that was less than or equal to half the assumed prevalence (1.25%).[3,4] We calculated a minimum sample size of 599. |
| Response: No | Study does not meet the required sample size. |
| Response: Unclear | Sample size not reported. |
| Notes | |
| Rationale for operationalization | A sample size calculation provides an objective numeric threshold to ease decision burden when critically appraising a study. Investigators may choose to change inputs for the sample size calculation depending on their preferences and goals for analysis. |
| Coding logic: *variables* (outcome) | This item may be completed using three *variables*:<br>● *Study defined sample size achieved* - a binary variable for whether a sample size calculation was provided by the study authors and whether the sample achieved the target size:<br> ● Calculation provided and sample achieved = Yes<br> ● Calculation provided and sample not achieved = No<br> ● Calculation not provided = No<br>● *Sample size* - a numeric variable reporting the included sample size<br>● *Sample size threshold* - a numeric variable linked to a sample size calculator<br>These three *variables* determine *Item C* (outcome) as follows: |

| | |
|---|---|
| | <ul><li>*Study defined sample size achieved* (Yes) = *Item C* (Yes)</li><li>*Sample size* > *Sample size threshold* = *Item C* (Yes)</li><li>*Study defined sample size achieved* (No) AND *Sample size* < *Sample size threshold* = *Item C* (No)</li></ul> |
| Risk of bias domain | Selection bias. |
| Principles guiding non-algorithmic risk of bias assessment and used to inform development of the algorithm | If this item is “No” then the study will not be marked at low risk of bias. This item can be the difference between moderate and high risk of bias. A value closer to the threshold could result in a study being at moderate risk of bias if all other criteria are met, whereas if the sample was very small then the study could be marked high risk of bias, even if all other items are met. |

| **Item D (JBI Item 5): Was data analysis conducted with sufficient coverage of the identified sample?** | |
|---|---|
| Response: Yes | The demographic characteristics of the sample were representative of the population in both the main and sub-group analyses.<br><br>Our assessment of seroprevalence studies evaluated reporting of age and sex. However, investigators may wish to consider an expanded list of potentially relevant characteristics. |
| Response: No | The demographic characteristics of the sample were not representative of the population in both the main and sub-group analyses. |
| Response: Unclear | Information is not provided about demographic characteristics of the sample. |
| Notes | |
| Rationale for operationalization | As highlighted by JBI guidance, not all subgroups of the population may respond at the same rate such that some subgroup analysis may no longer be representative of the population. It may be overwhelming to evaluate the degree to which the demographic characteristics of the sample are representative of the population in each analysis. Many demographic characteristics may be reported and the characteristics that are relevant for determining representativeness may vary by the type of disease being investigated. As such, this item is context specific and requires subjective judgement of the evaluators.<br><br>It may be reasonable to focus assessment on sex and age as they are key epidemiological variables and are often reported in disease prevalence studies. However, investigators may wish to consider an expanded list of potentially relevant characteristics. |
| Coding logic | Not applicable. This item requires subjective evaluation as to whether the sample is at least somewhat representative of the population in both the main and sub-group analyses. Investigators should review the demographic breakdown in the study and in each analysis, and review author comments on representativeness of the analyzed sample. |
| Risk of bias domain | Selection bias. |
| Principles guiding non-algorithmic risk of bias assessment and used to inform development of the algorithm | If the study used an appropriate sample frame, probability sampling methods were used, and had an adequate sample size, this item is usually marked as "Yes". Under these conditions, it is only marked as "No" if there is an obvious sub-group that is poorly represented - which is typically only clear if many sub-group estimates are provided. The study authors will typically identify this. |

<table>
<tr><th colspan="2">Item E (JBI item 6): Were valid methods used for the identification of the condition?</th></tr>
<tr><td>Response: Yes</td><td>The measurement test or a combination of measurement tests exceeded the threshold for adequate sensitivity and specificity according to leading health authorities and regulators (e.g., World Health Organization, Centers for Disease Control and Prevention, European Medicines Agency, U.S Food and Drug Administration, Health Canada) or used one of the following multi-test algorithms*:<br>● Two test algorithms using a commercial or in-house binding assay with confirmatory testing using virus neutralization assay irrespective of combined sensitivity/specificity<br>● Three test confirmatory approach during which two tests are used in parallel followed by a third confirmatory test for discordant results<br>● Three test series approach whereby positive is defined by at least two out of three tests being positive<br>● Three test parallel approach whereby positive is defined by at least one positive test and no negative tests<br><br>In our evaluation of seroprevalence studies the method was deemed valid if any of the following were true:<br>● The serological test used met the WHO Unity Study Criteria for serological tests: sensitivity minimum 90%, specificity minimum 97%[4]<br>● Multiple test algorithm with combined sensitivity ≥90% and combined specificity ≥97%<br>● Two test algorithms using a commercial or in-house binding assay with confirmatory testing using virus neutralization assay irrespective of combined sensitivity/specificity<br>● Three test algorithms designed as confirmatory, series, or parallel (described above)<br><br>*Users of this tool may wish to define their own thresholds for sensitivity and specificity or define other valid multi-test algorithms.</td></tr>
<tr><td>Response: No</td><td>The detection test does not exceed the threshold for adequate sensitivity and specificity according to leading health authorities and regulators (e.g., World Health Organization, Centers for Disease Control and Prevention, European Medicines Agency, U.S Food and Drug Administration, Health Canada) or did not use a valid multi-test algorithm (see above).<br><br>In evaluation of our sero-prevalence studies the method was deemed not valid if any of the following were true:<br>● The serological test used did not meet the WHO Unity Study Criteria for serological tests: sensitivity minimum 90%, specificity minimum 97%<br>● The combined sensitivity and specificity for the two-test algorithm was below the threshold<br>● Complex multiple test algorithms not using a valid multi-test algorithm (see above)</td></tr>
<tr><td>Response: Unclear</td><td>Test sensitivity and specificity not reported.</td></tr>
</table>

| Notes | |
|---|---|
| Rationale for operationalization | Use of objective numerical thresholds for test sensitivity and specificity may ease decision burden. Investigators may disagree on the source of the most appropriate thresholds. If there is disagreement on the thresholds considered adequate by different health authorities and regulators disagree, then the default selection should be the highest thresholds unless otherwise justified.<br><br>Multiple detection tests may be used in combination to improve overall detection. Sensitivity and specificity of the combined tests may not be reported as there may be a presumption of high accuracy. This was a common occurrence in seroprevalence testing during the COVID-19 pandemic. Use of decision rules for testing matrices provides an objective approach to ease decision burden. Three tests is the minimum number required for a tie-breaker evaluation in the event of mixed results arising from two tests. We identified strategies using three tests that may be considered sufficiently valid even in the absence of reported sensitivity and specificity: confirmatory, series, parallel. However, investigators may wish to disregard studies using detection tests without reported sensitivity and specificity. |
| Coding logic | This item may be completed using three *variables*:<br>● *Sensitivity* – numeric<br>● *Specificity* – numeric<br>● *Test strategy* – a categorical variable describing the strategy of the testing:<br>o Single test performance reported (sensitivity and specificity reported)<br>o Single test performance not reported (sensitivity and specificity not reported)<br>o Multi-test performance reported (sensitivity and specificity for the combined testing strategy was reported)<br>o Multi-test with confirming neutralization test (commercial or in-house binding assay with confirmatory testing using virus neutralization assay irrespective of combined sensitivity/specificity)<br>o Three test confirmatory (two tests in parallel followed by a third confirmatory test for discordant results)<br>o Three test series (three sequential tests whereby positive is at least two of three tests are positive)<br>o Three test parallel (three tests in parallel whereby positive is at least one positive test and no negative test in parallel testing with three tests)= Yes<br>o Multi-test strategy unclear<br><br>These three *variables* (outcome) determine *Item D* (outcome) as follows:<br>● *Test strategy* (Single test performance reported) AND *Sensitivity* ≥ threshold AND *Specificity* ≥ threshold = *Item D* (Yes)<br>● *Test strategy* (Multi-test performance reported) AND *Sensitivity* ≥ threshold AND *Specificity* ≥ threshold = *Item D* (Yes)<br>● *Test strategy* (Single test performance not reported) = Item D (No) |

| | |
|---|---|
| | ● *Test strategy* (Single test performance reported) AND *Sensitivity* < threshold AND *Specificity* <threshold = *Item D* (No)<br>● *Test strategy* (Multi-test performance reported) AND *Sensitivity* < threshold AND *Specificity* < threshold = *Item D* (No)<br>● *Testing strategy* (Multi-test with confirming neutralization test*) OR Testing strategy* (Three test confirmatory*)* OR *Testing strategy* (Three test series*) OR Testing strategy* (Three test parallel*)* = *Item D* (Yes)<br>● *Testing strategy* (Single test performance not reported) OR (Multi-test strategy unclear*)* = *Item D* (Unclear) |
| Risk of bias domain | Measurement bias |
| Principles guiding non-algorithmic risk of bias assessment and used to inform development of the algorithm | If this item is “No” then the study could still be marked at low risk of bias, if robust test adjustment is conducted and the other major items are met. |

| **Item F (JBI Item 7): Was the condition measured in a standard, reliable way for all participants?** | |
|---|---|
| Response: Yes | The same measurement test and procedures were used for all participants. |
| Response: No | Different measurement tests and procedures were used for participants. |
| Response: Unclear | No details were provided about which participants received which measurement tests. |
| Notes | |
| Rationale for operationalization | If the authors describe one detection test and set of procedures utilized for all participants, then standardization and reliability is presumed. |
| Coding logic | This item may be completed using one *variable*:<br>● *Consistent testing/procedures* – binary variable describing whether a single testing/procedure was utilized for all participants.<br><br>This one *variable* (outcome) determines *Item E* (outcome) as follows:<br>● *Consistent testing/procedures* (Yes) = *Item E* (Yes)<br>● *Consistent testing/procedures* (No) = *Item E* (No) |
| Risk of bias domain | Measurement bias. |
| Principles guiding non-algorithmic risk of bias assessment and used to inform development of the algorithm | This item is usually "Yes". Few studies use different tests on different participants and attempt to amalgamate the results. If different participants received different tests and the results were amalgamated, but the tests were all high-performing and robust test adjustment was used then this would not impact on the risk of bias. |

| **Item G (JBI Item 8): Was there appropriate adjustment for test characteristics?** | |
|---|---|
| Response: Yes | Provided statistical adjustment for test characteristics and the information necessary to determine the numerator, denominator, prevalence estimate, and confidence interval. |
| Response: No | Did not statistically adjust for test characteristics or did not provide the information necessary to determine the numerator, denominator, prevalence estimate, and confidence interval. If adjustment was not reported, then it was considered as having not been conducted. |
| Notes | |
| Rationale for operationalization | This item was derived from a single JBI item on statistical analysis. It was operationalized into two components – Item G focuses on test adjustment and Item H focuses on population characteristics - to account for the fact that studies may do one or the other and not both. There are different implications for risk of bias from failing to adjust for test characteristics and population characteristics, thus they were separated.<br><br>Test adjustment reduces measurement bias. A binary decision on whether statistical adjustment was completed eases decision burden when critically appraising the analytic methods of a study. However, different adjustment techniques are possible which may result in heterogeneity between studies. Investigators should have the option of conducting their own adjustment for the purpose of standardization. As such, this item was constructed to reduce decision burden by using a binary assessment of whether test adjustment occurred and use a binary assessment of whether key information was provided to enable adjustment of the prevalence estimates by the investigators. |
| Coding logic | This item may be completed using three *variables*:<br>● *Test adjustment* - binary variable on whether test adjustment was conducted<br>● *Sensitivity* - numeric<br>● *Specificity* - numeric<br><br>These three *variables* (outcome) determine *Item G* as follows:<br>● *Test adjustment* (Yes) = *Item G* (Yes)<br>● *Test adjustment* (Yes) AND *Sensitivity* ≥90% AND *Specificity* ≥97% = *Item G* (Yes)<br>● *Test adjustment* (No/Unclear) AND/OR *Sensitivity* <90% AND/OR *Specificity* <97% = *Item G* (No) |
| Risk of bias domain | Measurement bias. |
| Principles guiding non-algorithmic risk of bias assessment and used to inform development of the algorithm | If this item is “No” then the study will not be marked at low risk of bias. If the sample was representative, probability sampling methods were used, and it was a large sample size, then failure to population adjust would not impact on the risk of bias. |

| Item H (JBI item 8): Was there appropriate adjustment for population characteristics? | |
|---|---|
| Response: Yes | Provided statistical adjustment for population characteristics or the sample was representative of the population (probability sampling) and provided the information necessary to determine the numerator, denominator, prevalence estimate, and confidence interval. |
| Response: No | Did not statistically adjust for population characteristics and the sample was not likely representative of the population (non-probability sampling) or did not provide the information necessary to determine the numerator, denominator, prevalence estimate, and confidence interval. If adjustment was not reported, then it was considered as having not been conducted. |
| Notes | |
| Rationale for operationalization | This item was derived from a single JBI item on statistical analysis. It was operationalized into two components – Item G focuses on test adjustment and Item H focuses on population characteristics - to account for the fact that studies may do one or the other and not both. There are different implications for risk of bias from failing to adjust for test characteristics and population characteristics, thus they were separated.<br><br>Population adjustment reduces selection bias. A binary decision on whether statistical adjustment was completed eases decision burden when critically appraising the analytic methods of a study. However, failure to adjust for population may not impact on the risk of bias if the sample is already representative of the population. Furthermore, different adjustment techniques are possible which may result in heterogeneity between studies. Investigators should have the option of conducting their own adjustment for the purpose of standardization. As such, this item was constructed to reduce decision burden by using a binary assessment of whether population adjustment occurred, account for failures to adjust whereby a representative sample was still achieved and use a binary assessment of whether key information was provided to enable adjustment of the prevalence estimates by the investigators. |
| Coding logic | This item may be completed using four *variables*:<br>● *Population adjustment* - binary variable on whether population adjustment was conducted.<br>● *Item A* - Binary variable on whether the sample frame approximated the target population.<br>● *Item B* - Categorical variables on whether study participants were recruited in an appropriate way to reduce selection bias.<br>● *Item I* - Categorical variable on whether the response rate was adequate, and if not, whether the low response rate was managed appropriately |

| | |
|---|---|
| | These four *variables* (outcome) determine *Item H* as follows:<br>● *Population adjustment* (Yes) = *Item H* (Yes)<br>● *Population adjustment* (No) AND *Item A* (Yes) AND *Item B* (Yes) AND *Item I* (Yes) = *Item H* (Yes)<br>● *Population adjustment* (No) AND/OR *Item A* (No) AND/OR *Item B* (No/Unclear) AND *Item I* (No/Unclear) = *Item H* (No) |
| Risk of bias domain | Measurement bias. |
| Principles guiding non-algorithmic risk of bias assessment and used to inform development of the algorithm | If this is “No” then the study will not be marked at low risk of bias. Often this is unclear. If this is unclear but the other major items are met and there is a large sample size then typically we will still mark as low risk of bias. |

| **Item I (JBI Item 9): Was the response rate adequate, and if not, was the low response rate unlikely to introduce bias?** | |
|---|---|
| Response: Yes | Response rate was greater than or equal to 60% or the sample was representative of the target population.[5] |
| Response: No | Response rate was less than 60% and the sample was not representative of the target population. |
| Response: Unclear | The response rate was not provided and it was unclear if the sample was representative of the target population. |
| Notes | |
| Rationale for operationalization | Response rate is used as an indicator for the risk of selection bias. Investigators may consider different response rates as adequate. Use of an objective numeric response rate threshold may ease decision burden when critically appraising response rate. However, a response rate lower than any given threshold does not necessarily correspond to an increased risk of selection bias if the demographics of the sample match those of the target population. As such, this item was constructed to reduce decision burden by using a response rate threshold and account for potentially low response rates whereby a representative sample was still achieved. |
| Coding logic | This item can be completed using five *variables*:<br>● *Response rate* – numerical<br>● *Item A* - Binary variable on whether the sample frame approximated the target population.<br>● *Item B* - Categorical variables on whether study participants were recruited in an appropriate way to reduce selection bias.<br>● *Item C* - Binary variable on whether the sample size was adequate.<br>● *Item H* - Binary variable on whether population adjustment occurred.<br><br>These five variables (outcome) determine Item I (outcome) as follows:<br>● *Response rate* ≥60% OR *Item A* (Yes) AND *Item B* (Yes) AND *Item C* (Yes) AND *Item H* (Yes) = *Item I* (Yes)<br>● *Response rate* <60% AND *Item A* (No) OR *Item B* (No/Unclear) OR *Item C* (No) OR *Item H* (No) = *Item I* (No)<br>● *Response rate* (Missing) = *Item I* (Unclear) |
| Risk of bias domain | Selection bias. |
| Principles guiding non-algorithmic risk of bias assessment and used to inform development of the algorithm | If this is “No” then the study will not be marked at low risk of bias. Often this is unclear. If this is unclear but the other major items are met and there is a large sample size then this item may still be marked as low risk of bias. |

| Item J: Overall risk of bias | |
|---|---|
| Low | The estimates are very likely correct for the target population. To obtain a low risk of bias classification, all criteria must be met or departures from the criteria must be minimal and unlikely to impact on the validity and reliability of the prevalence estimate. Sampling biases must be limited (i.e., appropriate sample frame, probability sampling, adequate sample size, and statistical adjustment for population characteristics) and measurement biases must be limited (i.e., adequate sensitivity/specificity and/or adjustment for test performance). These include sample sizes that are just below the threshold when all other criteria are met, reporting only some of characteristics of the sample, test characteristics below the threshold but adjustment made for the test performance, and response rates that are just below the threshold in the context of probability-based sampling of an appropriate sampling frame with population weighted seroprevalence estimates. |
| Moderate | The estimates are likely correct for the target population. To obtain a moderate risk of bias classification, most criteria must be met and departures from the criteria are likely to have only a small impact on the validity and reliability of the prevalence estimates. Sampling must be somewhat representative of the target population (i.e., inappropriate sample frame but probability sampling, adequate sample size, and statistical adjustment for population characteristics) or measurement biases must be limited. |
| High | The estimates are not likely correct for the target population. To obtain a high risk of bias, many criteria must not be met or departures from criteria are likely to have a major impact on the validity and reliability of the prevalence estimates. Sampling not likely representative of the target population (i.e., non-probability sampling and inadequate sample size) and considerable measurement bias (i.e., poor sensitivity/specificity and lack of adjustment for test performance). |
| Unclear | There was insufficient information to assess the risk of bias. |
